# Safety and immunogenicity of PHH-1V as booster vaccination through the Omicron era: results from a phase IIb open-label extension study up to 6 months

**DOI:** 10.1101/2024.04.09.24305540

**Authors:** M.J. Lopez, M.M. Vazquez, M. Alvarez, J.R. Arribas, E. Arana-Arri, P. Muñoz, J. Navarro-Pérez, R. Ramos, J. Molto, S. Otero-Romero, I. Esteban, E. Aurrecoechea, R. Pomarol, M. Plana, R Perez-Caballero, L. Bernad, J.G. Prado, L. Riera-Sans, A. Soriano

## Abstract

**Background:** Phase IIb HIPRA-HH-2 study results showed that PHH-1V as first booster dose elicited a strong and sustained neutralising antibody response against various SARS-CoV-2 variants. Here, we report the safety and immunogenicity of a fourth booster dose of PHH-1V against the most prevalent Omicron SARS-CoV-2 variants in Spain.

**Methods:** The HIPRA-HH-2 open-label extension study (NCT05142553) evaluated the safety and immunogenicity of PHH-1V as a fourth booster dose in subjects aged ≥18 years and followed for 6 months. Subjects received a fourth dose of PHH-1V 6–12 months after a previous regime of either two doses of BNT162b2 plus a third dose of PHH-1V (Cohort 1) or three doses of BNT162b2 (Cohort 2). Primary regulatory endpoint evaluated the neutralisation titres (GMT) against Omicron BA.1 on Day 14 of PHH-1V used as fourth dose in Cohort 2 vs the BNT162b2 used as third dose in initial HIPRA-HH-2 study. The immunogenicity of PHH-1V as fourth dose was also investigated by GMTs against Beta, Delta, and Omicron BA.1, BA.4/5 and XBB.1.5 on Days 14, 98 and 182 post-immunisation in the overall study population and in Cohorts 1 and 2 versus baseline. Safety of the fourth dose was also assessed.

**Findings:** From September 2022, 288 subjects received PHH-1V as a fourth dose (Cohort 1 n=106; Cohort 2 n=182). A significant increase in neutralising antibodies against Omicron BA.1 subvariant at Day 14 was observed from the third homologous booster with mRNA vaccine compared to the fourth heterologous booster with PHH-1V (1739.02 vs 4049.01; GMT ratio 0.43 (95% CI: 0.28; 0.65; p-value < 0.0001). PHH-1V used as fourth booster induced a statistically significant increase in neutralising antibody titres 14 days after immunisation for all variants compared with baseline [GMFR on Day 14 (95%CI) was 6.96 (5.23, 9.25) for Beta variant; 6.27 (4.79, 8.22) for Delta variant; 9.21 (5.57, 15.21) for Omicron BA.1 variant; 11.80 (8.29, 16.80) for Omicron BA.4/5 variant and 5.22 (3.97, 6.87) for Omicron XBB.1.5 variant]. Titres remained significantly higher compared with baseline at 3 and 6 months post-vaccination. Cohort comparison revealed no significant differences at 14, 98 and 182 days post-vaccination. The most frequent adverse events were injection site pain (Cohort 1: 84.0%; Cohort 2: 77.5%) and fatigue (Cohort 1: 17.9%; Cohort 2: 29.1%). No subjects experienced severe COVID-19 infection.

**Interpretation:** The PHH-1V vaccine as a booster induced a potent and sustained neutralising antibody response against previous circulating Beta, Delta variants and Omicron BA.1, BA.4/5, and XBB.1.5 subvariants in subjects previously vaccinated with three doses regardless of previous regimen. These findings suggest that PHH-1V could be an appropriate strategy for upcoming heterologous vaccination campaigns.

**Funding:** HIPRA SCIENTIFIC, S.L.U (HIPRA), Spain.

**Research in context:** *Unmet needs:* Immunity against SARS-CoV-2 will continue to increase in the community through widespread vaccination and infection. Despite this, at the individual level, the humoral response against new variants is diminished in both vaccinated and infected individuals. Booster strategies have demonstrated a reduction in the risk of not only COVID-19 infection but also of long COVID-19 or persistent post-COVID manifestations. Furthermore, heterologous booster strategies for vaccination regimens offer broad neutralising responses. However, available evidence regarding new platforms beyond mRNA-based vaccines is currently limited.

*Evidence before this study:* The PHH-1V vaccine elicits high and long-lasting levels of neutralising antibodies against all COVID-19 variants studied, as well as a strong cellular immunity response, when used as a heterologous booster in previously vaccinated individuals receiving mRNA and viral vector vaccines. However, safety and immunogenicity data on a fourth booster dose of PHH-1V against the most prevalent Omicron variants in Spain were not available at the time of the study period.

*Added value of this study:* The PHH-1V dimeric adjuvanted vaccine delivered as a fourth booster dose can induce a potent and significant neutralising antibody response against previous circulating Beta, Delta variants and Omicron BA.1, BA.4/5, and also against XBB.1.5 subvariants from Day 14 through Day 182 compared with baseline regardless of the primary vaccination received (two doses of BNT162b2 plus a third dose of PHH-1V (Cohort 1) or three doses of BNT162b2 (Cohort 2)) and confirm the higher response of PHH-1V when used as a heterologous fourth-dose booster. This open-label extension study also demonstrated that PHH-1V is well tolerated and safe irrespective of the prior booster vaccination received.

*Implications of all the available evidence:* These data confirm the advantages of heterologous booster vaccination with PHH-1V and the broad-spectrum response of the PHH-1V vaccine against the different emerging variants of COVID-19, suggesting that PHH-1V could be an appropriate booster for upcoming heterologous vaccination campaigns.

## Introduction

Since the advent of severe acute respiratory syndrome coronavirus 2 (SARS-CoV-2)^1^ that emerged in Wuhan, China, in 2019,^2,3^ huge efforts had been made to develop multiple prophylactic strategies including vaccines. Nevertheless, severe COVID-19 disease accumulates up to 6.96 million deaths worldwide (10^th^ October 2023).^4,5^

At the population level, immunity against SARS-CoV-2 will increase through widespread immunisation and infection occurrence.^6^ Despite this, on an individual level, the humoral response against new variants is diminished for both SARS-CoV-2 vaccinated and infected individuals.^6^ Vaccines are needed that offer broad and long-lasting immunological protection and reduce the incidence of severe disease and related hospitalisations.^7^ In addition, the emergence of new variants for SARS-CoV-2 such as Omicron and its sub-variants,^9^ requires the adaptation of immunisation strategies^10^ by implementing effective vaccination regimens, preferably combining heterologous boosters. While viral vector vaccines use a modified version of a different virus as a vector to deliver protection,^11^ mRNA vaccines use genetically modified RNA to generate a protein that, in turn, elicits a safe immune response.^12^ Additionally, there are also adjuvanted protein-based subunit vaccines, such as PHH-1V, which represent a new generation of vaccines that elicit a safe and strong immune response targeted to key parts of the virus with a better reactogenicity profile than mRNA based vaccines,^13^ even in persons with a weakened immune system.^11^

Although primary vaccination offers good protection against severe disease, national and international studies using mRNA COVID-19 vaccines have shown a reduction in effectiveness in adults 3–6 months post-vaccination, especially in terms of infection rates.^8^ Consequently, most national vaccine strategies included additional booster doses with either the same vaccine type (homologous booster) or a different vaccine type (heterologous booster) as both approaches have proven to provide appropriate immunogenicity.^8^ In fact, booster strategies have demonstrated higher efficacy in reducing the risk of not only COVID-19 infection but also of long COVID-19 or persistent post-COVID manifestations in up to 73% of subjects after receiving three doses.^14^ In addition, the presence of adjuvants contributes to the induction and establishment of a sustained immune response, thus enhancing the overall magnitude and durability of immune response in the long-term.^15^

PHH-1V (BIMERVAX^®^; HIPRA, SPAIN) is a bivalent dimeric recombinant protein adjuvanted vaccine against SARS-CoV-2 and is based on a heterodimer protein comprising a recombinant receptor-binding domain (RBD) fusion from two SARS-CoV-2 variants, B.1.351 (Beta) and B.1.1.7 (Alpha). PHH-1V is indicated as a booster dose for active immunisation to prevent COVID-19 in people aged 16 years or older who have received a COVID-19 mRNA vaccine.^8,17–19^ On 30^th^ March 2023, the EMA recommended the approval of PHH-1V as a COVID-19 booster vaccine,^8,16,18,19^ on 1^st^ August 2023 it was authorised by the Medicines and Healthcare products Regulatory Agency (MHRA),^20^ and on 9^th^ October 2023 the World Health Organization (WHO) included PHH-1V in its list of pre-qualified vaccines.^21^

PHH-1V elicits high and long-lasting levels of neutralising antibodies against all COVID-19 variants studied, as well as a strong cellular immunity response, when used as a heterologous booster in previously vaccinated individuals with mRNA and viral vector vaccines.^8,17–19^

HIPRA-HH-2 (NCT05142553) study was a Phase IIb, randomised, double-blind, controlled, multicentre, non-inferiority clinical trial in 765 participants vaccinated against COVID-19 with BNT162b2 (tozinameran) at least 182 days prior to the administration of PHH-1V (n=513) or BNT162b2 (n=252) as first booster dose in 10 centres in Spain.^13,16^ Geometric mean titres (GMT) were studied after first booster dose and results indicated superiority of PHH-1V at Days 98 and 182 compared with BNT162b2 and non-inferiority at Days 14 and 28, depending on the variant evaluated.^13^ The overall frequency of adverse events (AEs) was significantly lower (p<0.05) among subjects who received the PHH-1V booster versus those receiving BNT162b2, with most AEs in both groups being mild.^13,16,22^ In addition, the PHH-1V vaccine was well tolerated and safe, regardless of vaccination history.^13,19,22^

The aim of our investigation was to evaluate the immunogenicity and safety of PHH-1V when administered as a fourth booster vaccination against SARS-CoV-2 variants of special interest in Spain. Due to the evolutive nature of the virus, herein we present results from the HIPRA-HH-2 open-label extension study and an additional analysis comprising new viral variants of interest in the Omicron era.

## Methods

### Study design and participants

HIPRA-HH-2, a phase IIb open-label extension study (NCT05142553) that evaluated the safety and immunogenicity of PHH-1V administered as a fourth booster dose in adult participants (≥18 years), started in September 2022 in 10 centres across Spain. The trial was conducted in accordance with the Declaration of Helsinki, the Good Clinical Practice guidelines, and national regulations. The study protocol was reviewed and approved by the Spanish Agency of Medicines and Medical Devices (AEMPS) as well as Independent Ethics Committee from the Hospital Clínic de Barcelona (HCB/2021/1110).

Study population were adult participants (≥18 years of age) who had completed 6 months on the HIPRA-HH-2 study and fulfilled the inclusion criteria to be enrolled in the extension phase of the study. Inclusion and exclusion criteria for the HIPRA-HH-2 study have been published previously.^13^ A fourth dose of PHH-1V was administered between 6–12 months to two cohorts of subjects which had received three previous doses of a SARS-CoV-2 vaccine based on different vaccination schemes: participants who had received prime vaccination with two doses of BNT162b2 plus a third dose of PHH-1V as booster (Cohort 1), and participants who received three doses of BNT162b2, two doses as prime vaccination and one as booster, (Cohort 2).^23^ Additionally, a group of participants from the community who matched the vaccination history of the HIPRA-HH-2 study and fulfilled the inclusion/exclusion criteria were recruited and included in Cohort 2 of the HIPRA-HH-2 extension phase to answer the primary objective (described below). Concomitant medications prohibited during the open-label study included anticoagulants, immunosuppressants and other immune-modifying treatments administered within 2 months before Day 0 and throughout the study.

Written informed consent was obtained from all participants before enrolment.

### Objectives

The primary objective of HIPRA-HH-2 extension study, which followed regulatory requirements, was to determine and compare the changes in immunogenicity measured by pseudovirion-based neutralisation assay (PBNA) against Omicron BA.1 subvariant at Day 14 post-fourth dose of PHH-1V in Cohort 2 vs BNT162b2 as post third dose in Cohort 2 from the initial HIPRA-HH-2 study. Secondary objectives were to determine and compare changes in immunogenicity (by PBNA) against Omicron BA.1, Omicron BA.4/5, and Beta and Delta variants at Days 14, 98 and 182 post-fourth dose of PHH-1V versus baseline, T-cell mediated response to the SARS-CoV-2 S protein at Days 14 and 182 post-fourth dose of PHH-1V in Cohort 2 and to assess the safety and tolerability of PHH-1V as a fourth dose. Additionally, due to clinical and public-health relevance of new emerging variants, a neutralising antibody analysis against Omicron XBB.1.5 was performed, although not included in the study protocol. Exploratory objectives included the assessment of reported COVID-19 severe infections occurring ≥14 days after booster and throughout the study.

### Study vaccine

PHH-1V was supplied in vials, each containing 10 ready-to-use doses of 0.5 mL (40 µg). There was no requirement to dilute or reconstitute the vaccine. PHH-1V was shipped to clinical sites and kept refrigerated at 2–8°C. Vials were not allowed to be frozen.

### Procedures and Outcomes

All eligible participants to receive a fourth dose of PHH-1V on Day 0 (open-label extension phase) were provided with a paper diary on Day 0 and returned to the site on Days 14. At day 0, 14, 98 and 182 (final visit) blood sample collection and safety follow-up were conducted.

Titres of neutralising antibodies were determined by the inhibitory concentration 50 (IC_50_, reported as reciprocal dilution) using a PBNA as described previously.^24^ The GMT and the geometrical mean fold rise (GMFR) for adjusted treatment were calculated.

The T-cell-mediated immune response against the SARS-CoV-2 spike glycoprotein and spike protein RBD sequence were assessed after the *in vitro* peptide stimulation of peripheral blood mononuclear cells (PBMC) from vaccinated participants followed by enzyme-linked immune absorbent spot (ELISpot) and intracellular cytokine staining (ICS) with several cytokines on baseline and at Days 14 and 182 post-fourth dose of PHH-1V in a subset of individuals from Cohort 2. Peptide pools of overlapping SARS-CoV-2 peptides, each encompassing the SARS-CoV-2 Protein S (two pools covering two different regions from spike S1 protein from Wuhan) or RBD domain (six peptides’ pools covering RBD domain of spike protein from Wuhan-Hu-1, Beta, Delta variants and Omicron BA.1, BA.2 and XBB.1.5 subvariants) were used (details of procedures have been published previously).^13^

As suggested by the regulatory agency, the primary study outcome measure was neutralisation antibody titre against Omicron BA.1, measured as the IC_50_ using PBNA and reported as log_10_ concentration for each individual sample and GMT, at Day 14 post-fourth dose with PHH-1V in Cohort 2 from the extension HIPRA-HH-2 study versus post-third dose with BNT162b2 in Cohort 2 from initial HIPRA-HH-2 study.

The secondary outcome measures were: neutralisation titres against Beta and Delta variants, and Omicron BA.1, BA.4/5, XBB.1.5 subvariants measured as the IC_50_ using PBNA and reported as log_10_ concentration for each individual sample and GMT, at Days 14, 98 and 182 post-fourth dose of PHH-1V versus baseline in both cohorts; the GMFR in neutralising antibody titres for all studied variants at Days 14, 98 and 182 post-fourth dose of PHH-1V versus baseline. T-cell-mediated immune response at baseline and Days 14 and 182 against SARS-CoV-2 Wuhan, Beta, Delta and Omicron variants.

Exploratory efficacy endpoint regarding severe COVID-19 infections occurrence from Day 14 post booster to Day 182 was assessed on the safety population.

Safety endpoints were solicited local and systemic reactions from the time of vaccination until 7 days post-vaccination self-reported in the subject diary provided to participants at study start. Safety was also assessed by recording treatment-emergent AEs (TEAEs) with onset on or after the administration of study treatment through Day 28. Treatment-emergent was defined as any AE with onset on or after the administration of study treatment through Day 28 or any event that was present at baseline but worsened in intensity or was subsequently considered drug related by the Investigator through the end of the study.

Serious AEs (SAEs), AEs of special interest (AESI) and medically attended AEs (MAAEs) were reported throughout the study. All AEs were coded using the Medical Dictionary for Regulatory Activities (MedDRA) Version 26.0 coding system.

### Statistical analysis

A sample size of 100 subjects was calculated to provide 90% power to detect non-inferiority of the fourth dose to the third dose for the primary endpoint Omicron BA.1 in Cohort 2 considering a 5% significance level, a non-inferiority margin of 1.5 and assuming a pooled standard deviation of 0.53. No sample size calculations were conducted for comparisons involving Cohort 1.

In this study, non-inferiority was to be determined if the upper bound of a 95% confidence interval (CI) surrounding the geometric mean ratio (GMT ratio) of the mean paired log-difference between the third dose and the fourth dose responses was below 1.5. If the upper bound of the 95% CI was also below 1, superiority was concluded.

To investigate the endpoints of neutralisation titres against viral subvariants, measured as IC_50_ by PBNA and reported as log_10_ concentration for each individual sample and GMT, at baseline and Days 14, 98 and 182 post-fourth dose of PHH-1V in Cohort 2 versus post-third dose, mixed models for repeated measures (MMRM) were used. Similarly, the comparison between the fourth dose in Cohort 1 versus the third dose in Cohort 2 was assessed using MMRM models as well. In these models, the log_10_-transformed neutralising antibody measurements were used as response variable while the study visit, the dose and the dose-by-visit interaction terms were used as fixed effects. The age group factor was considered a covariate and the site, and the subject-nested-to-site were introduced as random effects. A compound symmetry covariance matrix structure was used. The denominator degrees of freedom were computed using the Kenward-Roger method. Weights were applied to the model estimation to account for sample distributions across covariates. The weighted LS mean estimates for each treatment dose were presented with the associated standard errors and 95% CIs for all visits. The back-transformed treatment group LS mean estimates and difference in weighted LS means (GMT ratio) were also presented for all visits with the corresponding 95% CI and p-value. GMFR analyses were conducted with the log_10_-transformed post-baseline titre/baseline titre ratio using MMRM models as defined above.

Cellular immunogenicity analysis was analysed providing 2 values for each parameter, imputing 0 in the event of negative values. ELISpot data were provided as counts per million therefore the average value was divided by 1000000 for analysis. ICS data were provided as percentage therefore the data were divided by 100 for analysis. Also, MMRM were used for ELISpot data analysis and boxplot was used for graphical representation. For ICS data analysis, regression analysis was performed.

No formal hypothesis testing analysis of AE incidence rates was performed. Descriptive statistics were used for safety data reporting by cohorts and for the overall population.

## Results

In this open-label extension study, 301 subjects were screened of which 288 subjects were vaccinated with PHH-1V as a fourth dose and were distributed in two cohorts depending on the received previous vaccinations: Cohort 1 comprised a total of 106 subjects, all of them participants from previous HIPRA-HH-2 study, with two doses of BNT162b2 vaccine and one booster dose with PHH-1V; Cohort 2 included 182 subjects, 52 from the previous study and 130 subjects from the community (all having received two doses BNT162b2 vaccine and one additional dose BNT162b2 as booster) (Figure 1).

**Figure 1.**
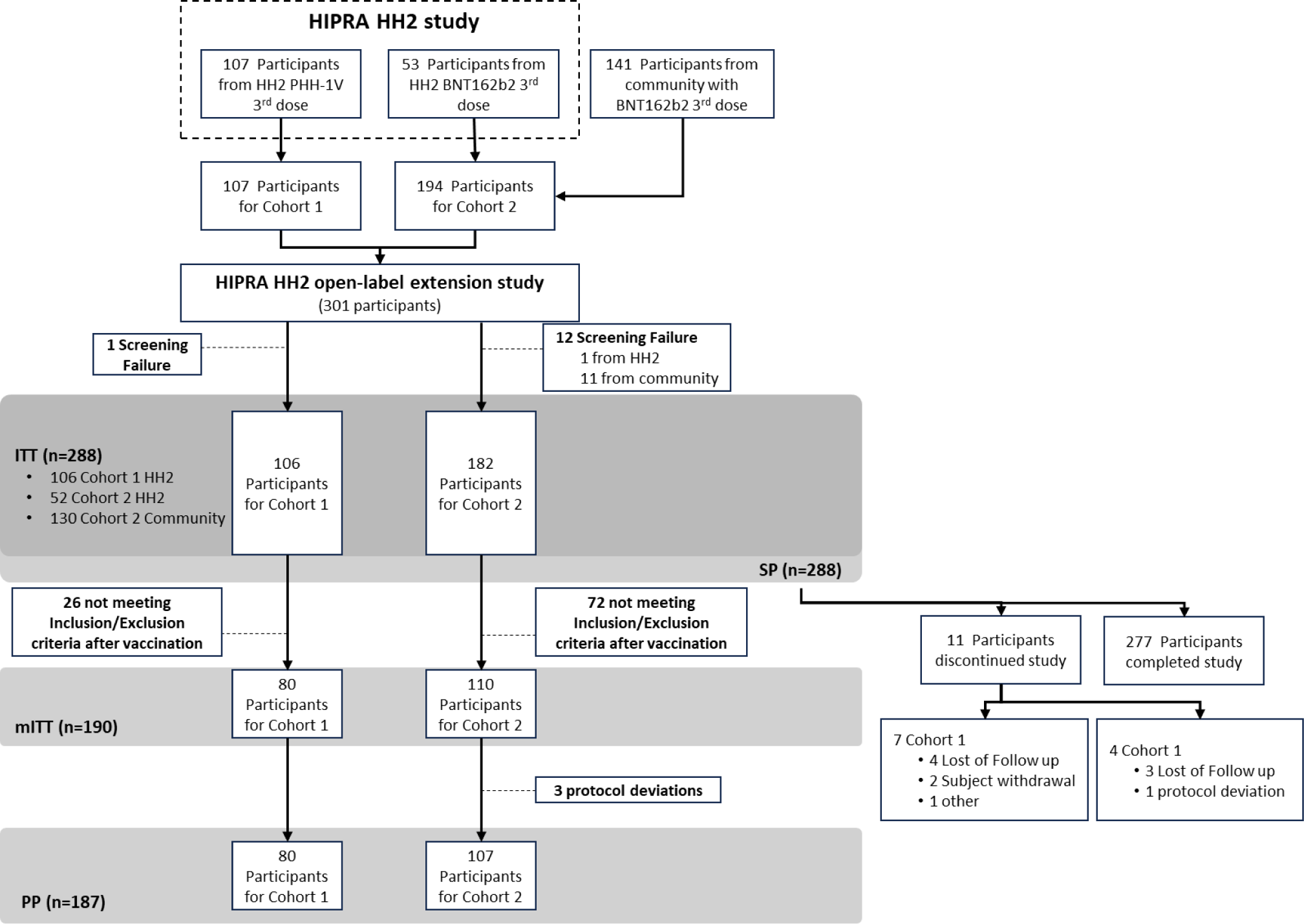
Patients disposition in HIPRA-HH-2 open-label extension study. Note: Subjects who tested positive for COVID-19 within 14 days of receiving study drug were excluded from the mITT.

Subjects’ demographics and baseline characteristics were balanced between cohorts and are shown in **Table 1**. For the overall population, mean age was 47.5 ± 14.86 years with 11.5% of the population aged 65 years or older. Baseline data from Cohort 2 community subjects reveals similar characteristics to those subjects already participating from the HIPRA-HH-2 study (data available in Table S1). Participant disposition showed that 11 (3.8%) subjects prematurely discontinued, 7 (6.6%) in Cohort 1 and 4 (2.2%) in Cohort 2. Reasons for early discontinuation were lost to follow-up (n=7, 2.4%), withdrawal (n=2, 0.7%), other (n=1, 0.3%) and protocol deviation (n=1, 0.3%). The mean study duration was 5.9 months (range: 1.0–6.5 months).

**Table 1.** Subjects’ demographics and baseline characteristics (safety population)

|  | **Cohort 1:**<br>**4<sup>th</sup> dose PHH-1V**<br>**(after 2 doses of**<br>**BNT162b2 + PHH**<br>**1V)**<br>**(n=106)** | **Cohort 2:**<br>**4<sup>th</sup> dose PHH-1V**<br>**(after 3 doses of**<br>**BNT162b2)**<br>**(n=182)** | **Overall**<br>**(N=288)** |
| --- | --- | --- | --- |
| **Mean age, years (SD)** | 49.0 (13.53) | 46.6 (15.55) | 47.5 (14.86) |
| **≥65 years of age, n (%)** | 12 (11.3) | 21 (11.5) | 33 (11.5) |
| **Sex, n (%)** |  |  |  |
| Male | 42 (39.6) | 73 (40.1) | 115 (39.9) |
| Female | 64 (60.4) | 109 (59.9) | 173 (60.1) |
| **Race, n (%)** |  |  |  |
| White | 106 (100) | 178 (97.8) | 284 (98.6) |
| Other | 0 | 4 (2.2) | 4 (1.4) |
| **Median body mass index at screening<sup>a</sup>,**<br>**kg/m<sup>2</sup> (range)** | 24.34<br>(18.03-39.78) | 24.92<br>(18.17-43.23) | 24.59<br>(18.03-43.23) |
| **Subjects with any medical history<sup>b</sup>,**<br>**n (%)** | 85 (80.2) | 130 (71.4) | 215 (74.7) |
| Hypertension | 24 (22.6) | 26 (14.3) | 50 (17.4) |
| Menopause | 14 (13.2) | 24 (13.2) | 38 (13.2) |
| Dyslipidaemia | 9 (8.5) | 9 (4.9) | 18 (6.3) |
| Hypercholesterolaemia | 4 (3.8) | 13 (7.1) | 17 (5.9) |
| Hypothyroidism | 6 (5.7) | 8 (4.4) | 14 (4.9) |
| Post menopause | 7 (6.6) | 6 (3.3) | 13 (4.5) |
| Asthma | 4 (3.8) | 7 (3.8) | 11 (3.8) |
| Type 2 diabetes mellitus | 2 (1.9) | 8 (4.4) | 10 (3.5) |

<sup>a</sup>Body mass index was not available for 1 subject.
<sup>b</sup>Medical history events were coded using the MedDRA Dictionary, version 26.0, and only those with an incidence of >3% are included.
N: number of patients.

### Immunogenicity of PHH-1V as heterologous booster

The extension study met its primary endpoint; a significant increase in neutralising antibodies at Day 14 post-administration of PHH-1V was observed from 1739.02 (95% CI: 18.30; 43.64) for the third dose with homologous booster to 4049.01 (95% CI: 2795.39; 5864.84) for the fourth dose as heterologous booster with PHH-1V (GMT ratio third dose vs fourth dose of 0.43 (95% CI: 0.28; 0.65); p value < 0.0001) against SARS-CoV-2 Omicron BA.1 variant (**Figure 2**). No significant differences were found at Day 14 between extension study Cohorts 1 and 2 with titres of 3521.41 (95% CI: 382.17, 822.56) and 3912.01 (95% CI: 2707.46, 5652.49), respectively (GMT ratio of 0.90 (95% CI: 0.59, 1.38); p value = 0.6316).

**Figure 2.**
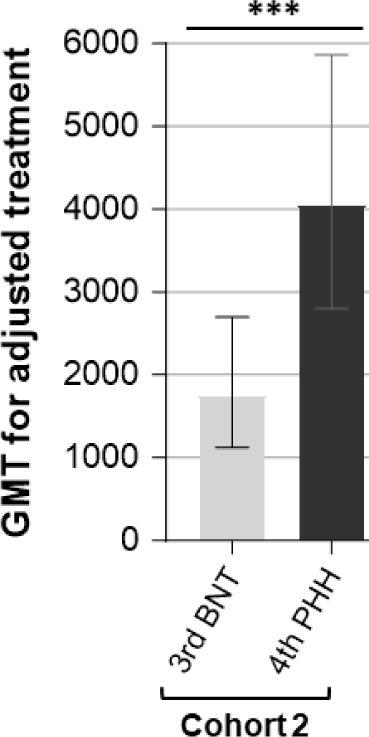
Comparison of neutralising antibody titre against SARS-CoV-2 Omicron BA.1 variant by PBNA. Representation of mean GMT for adjusted treatment and 95% CI at Day 14 post-vaccination for third dose with BNT162b2 vaccine from HIPRA-HH-2 study (third BNT; *light grey column*) and fourth dose with PHH-1V from HIPRA-HH-2 extension study (fourth PHH; dark grey column). CI: confidence interval; GMT: Geometric mean titre; PBNA: pseudovirion-based neutralization assay. *** p< 0.0001.

### Immunogenicity of PHH-1V booster dose

Humoral immune response of PHH-1V booster (fourth dose) against Beta, Delta, Omicron BA.1, Omicron BA 4/5 and Omicron XBB.1.5 variants were determined by neutralising antibody GMTs and GMFRs for modified intention-to-treat (mITT) population (n= 190; **Figure 3 and Table S2**). Fourth dose with PHH-1V induced a statistically significant increase in neutralising antibody titre on Day 14 for all variants compared with baseline titres. GMFRs (95% CI) at Day 14 were 6.96 (5.23, 9.25) for Beta variant, 6.27 (4.79, 8.22) for Delta variant, 9.21 (5.57, 15.21) for Omicron BA.1 variant, 11.80 (8.29, 16.80) for Omicron BA.4/5 variant and 5.22 (3.97, 6.87) for Omicron XBB.1.5 variant. Neutralising antibody titre results over time (**Figure 3 and Table S2**) revealed a decline in neutralising antibodies titres for all variants at 3 and 6 months after receiving the booster immunisation but titres remained significantly higher compared with baseline levels for the overall mITT population.

**Figure 3.**
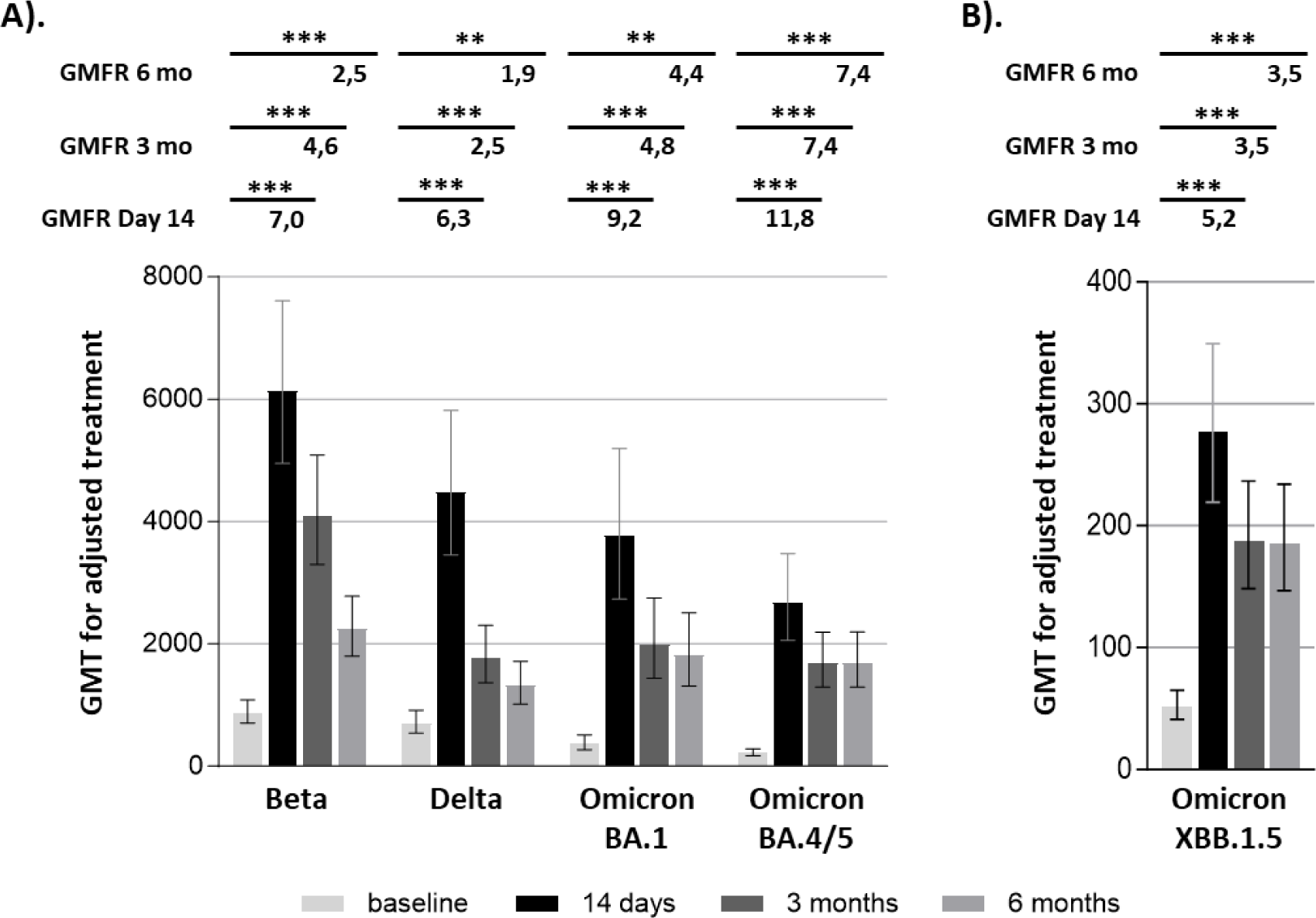
Neutralising antibody levels against SARS-CoV-2 variants by PBNA over time. Representation of mean GMT for adjusted treatment with 95% CI (columns) and mean GMFR from baseline (upper numbers) for all participants from mITT population treated with fourth dose of PHH-1V (n= 190) against SARS-CoV-2 Beta, Delta, Omicron BA.1, Omicron BA.4/5 (**A**) and Omicron XBB.1.5 variants (**B**) at baseline (*light grey*), Day 14 (*black*), 3 months (*dark grey*) and 6 months (*grey)* post-dose. Subjects who reported COVID-19 infections were excluded from the reported day onwards. CI: confidence interval; GMFR: Geometric mean fold rise; GMT: Geometric mean titre; mITT: modified intention-to-treat population; PBNA: pseudovirion-based neutralization assay. ***p<0.0001; **p<0.001.

Cohort analysis comparison of neutralising antibodies titres of the fourth dose with PHH-1V (Cohort 1: Homologous fourth dose, -n=80; Cohort 2: Heterologous fourth dose, n=110) were similar across subgroups for any variant at Day 14, 3 months and 6 months after the PHH-1V booster (**Table S3**). Statistically significant increases were observed from baseline following the fourth dose for all pre-specified variants with the exception of Omicron XBB.1.5 (**Table S3**).

SARS-CoV-2-specific T-cell responses after PHH-1V heterologous booster were evaluated in 15 participants from Cohort 2 by ELISpot and intracellular cytokine staining from PBMC at baseline, and at Day 14 and 6 months after dose. *In vitro* re-stimulation of PBMC from participants with SARS-CoV-2-derived peptide pools induced a significant IFN-γ T-cell response at Day 14 that persisted for at least 6 months (**Figure 4**). PHH-1V fourth dose significantly increased (p< 0.0001) the number of IFN-γ^+^ spot forming cells that responded to the *in vitro* PBMC re-stimulation with RBD (Wuhan, Beta, Delta, Omicron BA.1, Omicron BA.2 and Omicron XBB.1.5) and Spike A peptides pools on Day 14 compared with baseline (**Figure 4**). At 6 months after the fourth dose, increases compared with baseline were still significant for the number of IFN-γ^+^ spot forming cells that responded to the RBD Omicron BA.1 and Omicron XBB.1.5 variants (p< 0.05), and trends (0.05 < p < 0.1) to higher values were observed in response to the stimulation with RBD Wuhan and Omicron BA.2. Results from ELISpot against Omicron XBB.1.5 variant demonstrated that a fourth dose of PHH-1V elicited a higher IFN-γ^+^ T-cell response at Days 14 (78.70 IFN-γ^+^ spots/10^6^ PBMCs [range: 10.63–216.25]) and 182 after the booster (50.67 IFN-γ^+^ spots/10^6^ PBMCs [range: 3.75– 135.00]) compared with baseline (19.64 IFN-γ^+^ spots/10^6^ PBMCs [range: 0.00–50.00]).

**Figure 4.**
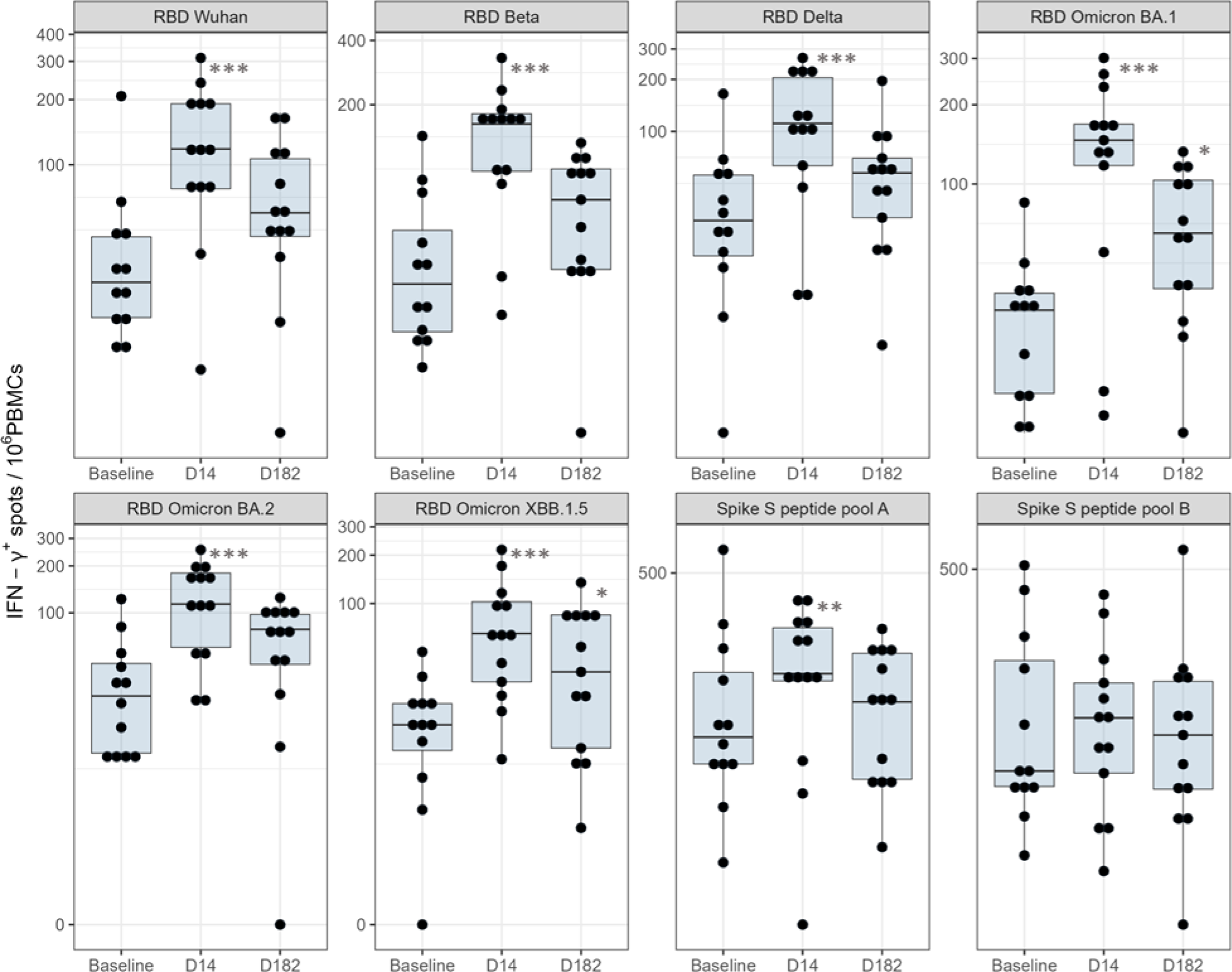
Total IFN-γ producing T cells upon PBMC re-stimulation with SARS-CoV-2 derived peptide pools by ELISpot. Frequencies of INF-γ^+^ cells determined by ELISpot assay in re-stimulated PBMC from participants with PHH-1V heterologous booster (cohort 2; n=15) isolated before (baseline), 2 weeks (Day 14) and 6 months (Day 182) after fourth dose and re-stimulated with RBD Wuhan D614G, RBD 1.351 (Beta), RBD B.1.617.2 (Delta), RBD Omicron BA.1, Omicron BA.2, XBB1.5 and Spike (A and B) peptides pools. ***p< 0.0001, **p< 0.01, *p< 0.05.

In the same subpopulation of 15 subjects from Cohort 2, the cellular immune response was analysed by ICS on CD4^+^ or CD8^+^ T-cells (**Figure 5**). ICS results showed that the stimulation of PBMC with the RBD (Wuhan, Beta, Delta, Omicron BA.1 and Omicron BA.2 variants) and Spike A peptide pools significantly induced a higher activation of CD4^+^ IFN-γ^+^ T-cells at Day 14 compared with baseline. The percentage of CD4^+^ IFN-γ^+^ T-cells responding in vitro to the RBD and Spike A stimulus decreased on Day 182 and no significant differences were observed in any T-cell responses at this time point compared with baseline (**Figure 5A**). Although a trend to higher values of CD8+ IFN-γ^+^ T-cells at Day 14 compared with baseline were observed after stimulation with RBD (Beta, Delta, Omicron BA.1 and Omicron BA.2 variants) and Spike A peptide pools, no statistically significant differences were observed (**Figure 5B**). No clear induction was seen for ICS of IL-4 or IL-2 (data not shown).

**Figure 5.**
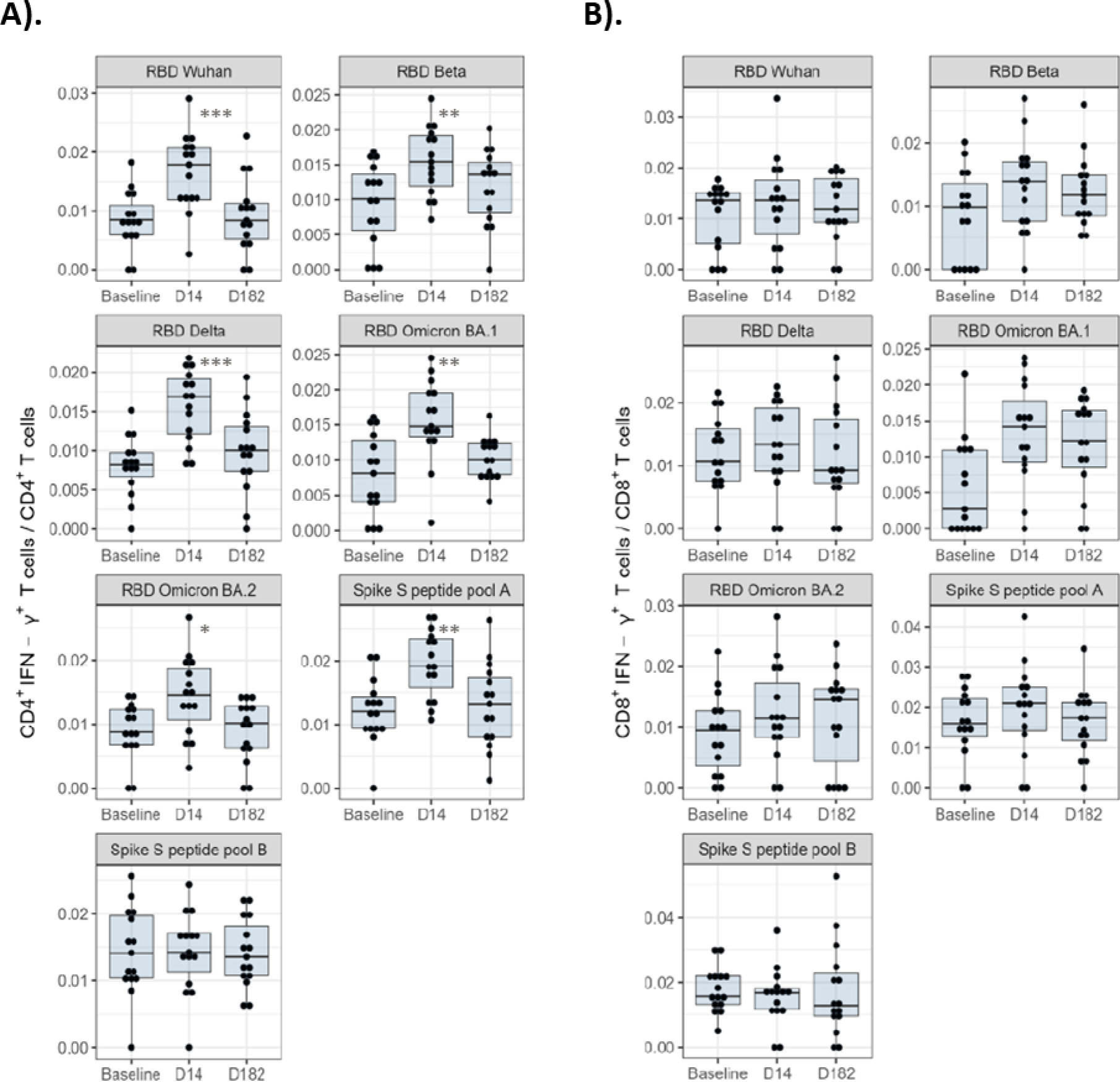
IFN-γ producing CD4^+^ and CD8^+^ T-cells upon PBMC re-stimulation with SARS-CoV-2-derived peptide pools by ICS. The frequencies of INF-γ expressing CD4^+^ T-cells (A) or CD8^+^ T-cells (B) are shown. PBMC were isolated from PHH-1V heterologous booster participants (Cohort 2; n= 15) before the immunisation (baseline), two weeks (Day 14) and 6 months (Day 182) after the fourth dose with PHH-1V, stimulated with RBD Wuhan D614G, RBD 1.351 (Beta), RBD B.1.617.2 (Delta), RBD Omicron BA.1, Omicron BA.2 and Spike (A and B) peptides pools, respectively. The cytokine expression in medium-stimulated PBMC was considered as the background value and subtracted from peptide-specific responses. ***p< 0.0001, **p< 0.001, *p< 0.01.

### Efficacy

At the end of the study, 36 subjects experienced non-severe COVID-19 infections (15 (14.2%) in Cohort 1 and 21 (11.5%) in Cohort 2). No subject experienced a severe COVID-19 infection, was hospitalized, admitted to the ICU, or died due to COVID-19 (**Table S4**).

### Safety

A total of 859 TEAEs were reported in 246 (85.4%) subjects, including 307 TEAEs in 92 (86.8%) subjects in Cohort 1 and 552 TEAEs in 154 (84.6%) subjects in Cohort 2. No subjects experienced a serious TEAE or a TEAE leading to death. Overall, 759 TEAEs were reported as mild in intensity in 188 (65.3%) subjects, 91 TEAEs were reported as moderate in intensity in 51 (17.7%) subjects, and 9 TEAEs were reported as severe in intensity in 7 (2.4%) subjects. The most frequent AEs were injection site pain (Cohort 1: 84.0%; Cohort 2: 77.5%) and fatigue (Cohort 1: 17.9%; Cohort 2: 29.1%) (**Table 2**). COVID-19 was reported as a TEAE in 4 (3.8%) subjects from Cohort 1 and 2 (1.1%) subjects from Cohort 2.

**Table 2.**
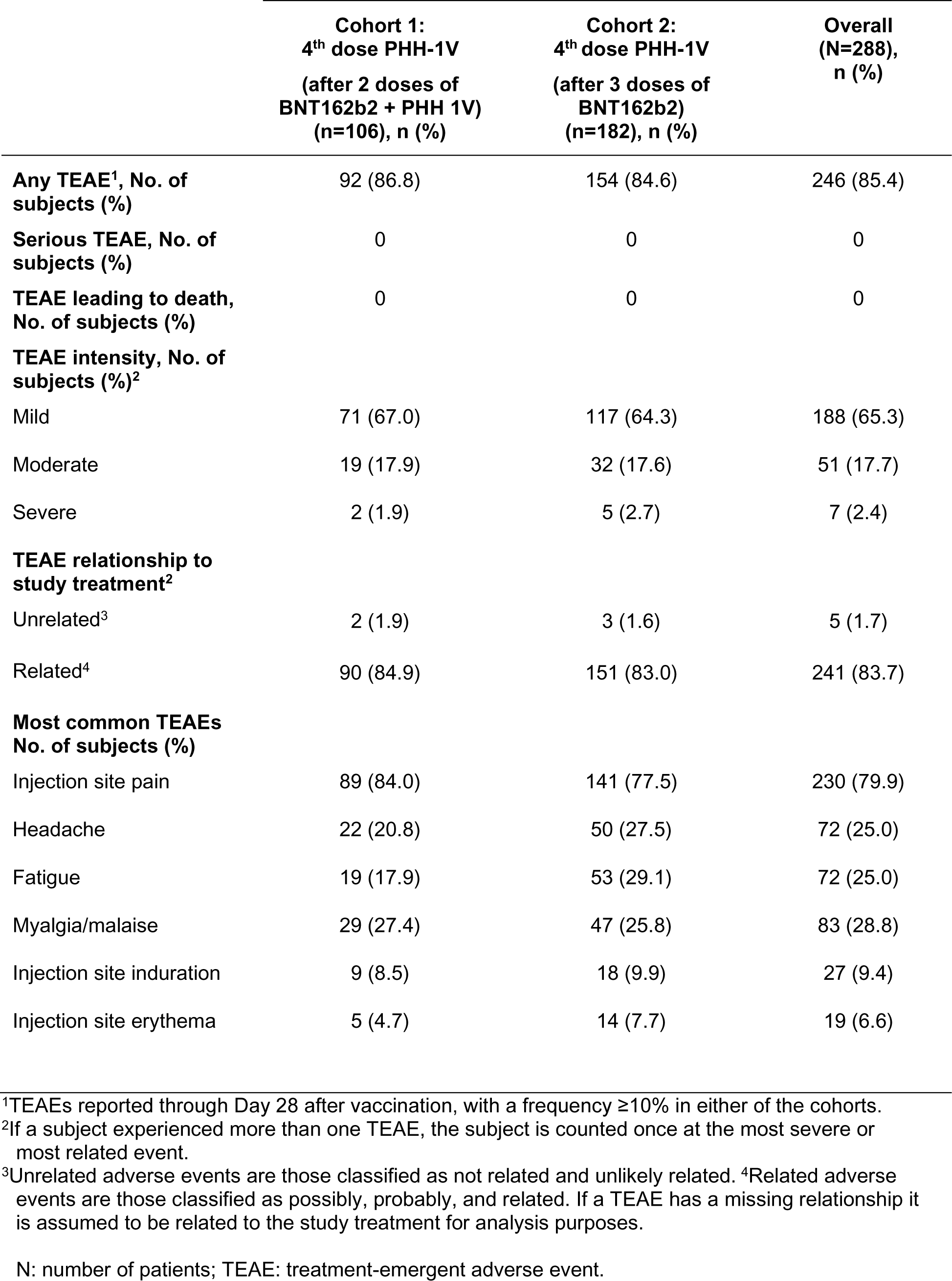
Frequency of TEAEs up to Day 28 by treatment group among the safety population of HIPRA-HH-2 extension study

|  | **Cohort 1:**<br>**4<sup>th</sup> dose PHH-1V**<br>**(after 2 doses of**<br>**BNT162b2 + PHH 1V)**<br>**(n=106), n (%)** | **Cohort 2:**<br>**4<sup>th</sup> dose PHH-1V**<br>**(after 3 doses of**<br>**BNT162b2)**<br>**(n=182), n (%)** | **Overall**<br>**(N=288),**<br>**n (%)** |
| --- | --- | --- | --- |
| **Any TEAE<sup>1</sup>, No. of subjects (%)** | 92 (86.8) | 154 (84.6) | 246 (85.4) |
| **Serious TEAE, No. of subjects (%)** | 0 | 0 | 0 |
| **TEAE leading to death, No. of subjects (%)** | 0 | 0 | 0 |
| **TEAE intensity, No. of subjects (%)<sup>2</sup>** |  |  |  |
| Mild | 71 (67.0) | 117 (64.3) | 188 (65.3) |
| Moderate | 19 (17.9) | 32 (17.6) | 51 (17.7) |
| Severe | 2 (1.9) | 5 (2.7) | 7 (2.4) |
| **TEAE relationship to study treatment<sup>2</sup>** |  |  |  |
| Unrelated<sup>3</sup> | 2 (1.9) | 3 (1.6) | 5 (1.7) |
| Related<sup>4</sup> | 90 (84.9) | 151 (83.0) | 241 (83.7) |
| **Most common TEAEs**<br>**No. of subjects (%)** |  |  |  |
| Injection site pain | 89 (84.0) | 141 (77.5) | 230 (79.9) |
| Headache | 22 (20.8) | 50 (27.5) | 72 (25.0) |
| Fatigue | 19 (17.9) | 53 (29.1) | 72 (25.0) |
| Myalgia/malaise | 29 (27.4) | 47 (25.8) | 83 (28.8) |
| Injection site induration | 9 (8.5) | 18 (9.9) | 27 (9.4) |
| Injection site erythema | 5 (4.7) | 14 (7.7) | 19 (6.6) |

<sup>1</sup>TEAEs reported through Day 28 after vaccination, with a frequency ≥10% in either of the cohorts.
<sup>2</sup>If a subject experienced more than one TEAE, the subject is counted once at the most severe or most related event.
<sup>3</sup>Unrelated adverse events are those classified as not related and unlikely related. <sup>4</sup>Related adverse events are those classified as possibly, probably, and related. If a TEAE has a missing relationship it is assumed to be related to the study treatment for analysis purposes.
N: number of patients; TEAE: treatment-emergent adverse event.

One SAE was reported in 1 (0.3%) subject from Cohort 1 during the extension phase. This SAE of thermal burn was assessed as unrelated to the study drug by the Investigator and Sponsor.

## Discussion

In the current endemic situation with emerging Omicron variants and waning protection, it is necessary to have booster vaccines with a broad response capability (breadth) to the different variants that also provide a long-lasting immune response. In addition, their inclusion in vaccination programmes for persons at risk of suffering severe disease (such as those over 60 years old, those with underlying diseases or immunocompromised persons) is crucial. The usefulness of booster doses against SARS-CoV-2 to prevent long COVID-19 has also been recently demonstrated.^14,24^ However, SARS-CoV-2 evolution, mainly driven by mutations in RBD allowing viral escape from neutralising antibodies, is responsible for limited vaccine efficacy, as has been observed with the emergence of the Omicron variant despite two-dose vaccination of WH1.^25^ Heterologous, rather than homologous, vaccination provides additional support for a mix-and-match approach^26^ and may provide more opportunities to accelerate the global vaccination campaigns. Currently available evidence comes from studies that include vaccines based on mRNA platforms and existing mRNA vaccines have not demonstrated sufficient immunity duration (up to 6 months), although new generation vaccines with self-amplifying mRNA appear to be a new option to improve the immunogenicity duration being non-inferior to BNT162b2.^27^ Recently, subtle differences in the mechanisms by which the different vaccine platforms (mRNA-, adenoviral vector-and recombinant protein-based) elicit immune responses have emerged:^28^ the two mRNA vaccines approved to date showed efficacy after dose one by means of non-neutralising antibodies and moderate Th1 responses while adenovirus vaccines elicited polyfunctional antibodies and potent T cell responses.^28^

The PHH-1V vaccine has proven its value as a booster in people with different primo-vaccination schedules (mRNA and/or adenovirus) as it is able to generate a potent, broad, and long-lasting immune response.^13,22^ The reason for this might be the result of several factors, including its dimeric structure and the use of adjuvant. PHH-1V is a bivalent antigen that allows the spike RBD sequence of two different SARS-CoV-2 variants to be contained in a single heterodimeric molecule. This heterodimer structure allows the booster-induced immune response to focus on an important region of the virus involved in target cell binding. The RBD sequence is immunodominant and accounts for 90% of serum neutralizing activity.^29^ Furthermore, the adjuvant enhances and induces an earlier, more robust and long-lasting immune response against the recombinant RBD heterodimer.^8^ The fact that PHH-1V contains RBD sequences is very relevant since it is the main target of neutralising antibodies (90% of the neutralising activity is associated with this region).^29^

Results of our open-label extension study are consistent with previous findings^13,22^ and demonstrate that PHH-1V is a robust immunogenic booster. The study met its primary objective and showed a significant increase in humoral immune response against the SARS-CoV-2 Omicron BA.1 variant using a heterologous booster with PHH-1V versus a homologous booster. The PHH-1V booster dose after a primary immunisation either in participants with three doses of BNT162b2 or those who received two doses of BNT162b2 and one dose of PHH-1V elicited an immune response and a cross-reactivity among all subvariants tested. Neutralising antibody titres were superior for a fourth dose of PHH-1V compared with baseline for the immune response on Days 14, 98 and 182 days irrespective of treatment cohort. Secondary endpoints revealed that the booster with PHH-1V induced a specific IFN-γ^+^ T-cell response against RBD peptides of SARS-CoV-2 Omicron BA.1, BA.2 and XBB.1.5 subvariants, showing the cross-reactivity of the cellular immune response induced by the PHH-1V vaccine. Moreover, the ICS results suggest that the booster with PHH-1V induced a Th1-biased cell response against all SARS-CoV-2 RBD variants tested on Day 14. However, the cellular immunity induced by the PHH-1V booster was not detected using ICS 6 months after immunisation, although T-cell responses were detected by ELISpot after in vitro re-stimulation with RBD peptides, which could be due to differences in sensitivity of the assays. The T-cell immune response generates memory T cells specific to the antigen that evoked the response, a key success factor for a vaccine.

These results are consistent with the observations of the HIPRA-HH-2 study and confirm the induction of strong humoral and cellular immunogenicity by PHH-1V, used both as a third dose^20^ and here as a fourth dose.

Safety data on PHH-1V as a fourth dose were consistent with the previously reported data following a third dose of PHH-1V.^13,22,30^ Only one serious AE was reported in 1 subject, which was assessed as unrelated to the study drug. There were no cases of severe COVID-19 infection in the open-label extension part of the study. These findings indicate that PHH-1V administered as a fourth dose provided protection against severe, life-threatening, and fatal forms of SARS-CoV-2 infection. Safety data also demonstrated the low reactogenicity of the vaccine, particularly the low incidence of fever. PHH-1V administered either as a fourth or a third dose demonstrated a good safety profile.^13,22^

An additional key benefit of adjuvant-based vaccines, such as PHH-1V, is that the inclusion of the adjuvant amplifies the immune response and positively impacts both the duration of immunity (humoral/cellular) and the breadth of the response. The value of adjuvanted protein vaccine boosting reported here has also been demonstrated elsewhere. For example, the monovalent beta-adjuvanted MVB.1.351 vaccine resulted in a higher neutralising antibody response against the original strain, the Beta variant and the Delta and Omicron BA.1 variants than those observed with the mRNA vaccine BNT162b2 and the MVD614 formulation.^31^ A two-dose regimen of the NVX-CoV2373 vaccine, which contains the full-length spike glycoprotein of the prototype strain plus Matrix-M adjuvant, administered to adults conferred 89.7% (95% CI 80.2 to 94.6) protection against SARS-CoV-2 infection and high efficacy against the B.1.1.7 variant.^28^ A post-hoc analysis showed an efficacy against the B.1.1.7 (or alpha) variant of 86.3% (95% CI 71.3 to 93.5) and against the non-B.1.1.7 variant of 96.4% (95% CI 73.8 to 99.5).^28^ From a practical perspective, protein-based vaccines offer convenient logistic features. These vaccines can be refrigerated as a ready-to-use formulation that does not require reconstitution prior to use, making distribution easier.^8^ These positive practical aspects make the PHH-1V vaccine convenient to use for forthcoming vaccination campaigns. In contrast, storage and transport of mRNA vaccines require rigorous temperature control, making them almost impossible to use in some countries.^32^ Additionally, it should be noted that immunocompromised patients and those on immunosuppressant therapies were excluded from mRNA vaccine trials because the neutralising antibodies resulting from vaccination can elicit an immunological cascade that may further deteriorate the general health of these individuals and increase the risk of viral infection.^32^

The immune response shown by PHH-1V booster against Wuhan, Beta, Delta and Omicron variants (BA.1; BA.4/5) in this extension study was comparable with the response triggered by PHH-1V in a previous study, while the XBB.1.5 neutralising antibody titres were lower, which was consistent with the results of other vaccine boosters and with the neutralising antibodies raised by a natural infection. It should be noted that both the vaccine (PHH-1V) and natural infection elicited a more discrete humoral response against XBB.1.5 compared with the other subvariants.^33^ It should also be pointed out that the evaluation of efficacy was extremely positive in terms of the incidence of COVID cases with a total of 36 cases (12.5%) being recorded, none of which were severe, during the 6-month study period when there was a high prevalence of BQ.1 and XBB.1.5 variants.^33^ The response to XBB.1.5 seems to confer sufficient protection against severe disease.^33^

In conclusion, the PHH-1V vaccine delivered as a booster dose induced a potent and significant neutralising antibody response against all studied Omicron variants up to XBB.1.5 via the same mechanism demonstrated previously for variants such as Wuhan, Beta and Delta.^30^ This confirms the broad-spectrum response of PHH-1V against the different emerging variants of COVID-19, including the XBB.1.5 subvariant although this is not as strong. In addition, studies with long follow-up (6–12 months) demonstrated that PHH-1V provides durable immune responses.^13,22^ This open-label extension study also demonstrated that PHH-1V was well tolerated and safe regardless of the primary vaccination received or previous SARS-CoV-2 infection. These findings suggest that PHH-1V could be an appropriate strategy for implementing upcoming heterologous vaccination campaigns.

### Contributions

All authors had full access to all study data, interpreted the data, provided critical conceptual input, critically reviewed and revised the manuscript and approved the decision to submit for publication. JML, MMV, MA, JRA, EAA, PM, JNP, RR, JM, SOR and AS are PI from the participating hospitals and listed in order of subject contribution. MP, IE, EA, RP, JGP, RPC, LB and IE were involved specifically in cellular immune response assay development, data generation and data analysis. LRS was involved on data analysis, manuscript writing and coordination.

### Data sharing statement

All data relevant to the study are included in the article or uploaded as supplementary information. Further data are available from the authors upon reasonable request and with permission of HIPRA S.A.

## Data Availability

All data produced in the present study are available upon reasonable request to the corresponding author

## Acknowledgments

Medical writing support was provided by Marta Morros (Adelphy Targis S.L., Barcelona, Spain) during the preparation of this paper and funded by HIPRA SCIENTIFIC, S.L.U.

Special thanks to all HIPRA members involved on study management, data management, pharmacovigilance, biostatistics analysis, quality.

We acknowledge the contribution of Ruth Peña for technical assistance with sample management and ELISpot and Felipe for data base generation. Moreover, we are indebted to the HCB-IDIBAPS Biobank, integrated in the Spanish National Biobanks Network, for the biological human samples and data procurement.

We especially acknowledge the following members of Veristat, who contributed to the success of this trial. The following were responsible for study management, biostatistics, medical monitoring, data management, and database programming of the study: Lubia Álvarez, MD, Robin Bliss, PhD, Judith Oribe, Emma Albacar, MPH, Nancy Hsieh, MPH, Marcela Cancino, MSc, Rachel Smith, Montse Barcelo, MD, Mariska van der Heijden, MSc, Amy Booth, Edmund Chiu and Rodney Sleith, MS, Avani Patel, Atalah Haun, MD and Cesar Wong, MD.

## Supplementary data

**TS1.** Subjects’ demographics.

TS1. Subjects' demographics.
|  |  | Cohort 1 | Cohort 2 |  | Overall |
| --- | --- | --- | --- | --- | --- |
|  |  |  | Part A | Community |  |
|  | n | 106 | 52 | 130 | 288 |
| Age | Mean age, years (SD) | 49.0 (13.53) | 46.1 (14.32) | 46.8 (16.06) | 47.5 |
|  | ≥65 years of age, n (%) | 12 (11.3) | 3 (5.8) | 18 (13.8) | 33 (11.5) |
| Sex; n (%) | Male | 42 (39.6) | 15 (28.8) | 58 (44.6) | 115 (39.9) |
|  | Female | 64 (60.4) | 37 (71.2) | 72 (55.4) | 173 (60.1) |
| Race; n (%) | White | 106 (100.0) | 51 (98.1) | 127 (97.7) | 284 (98.6) |
|  | Other | 0 | 1 (1.9) | 3 (2.3) | 4 (1.4) |
| Weight | Median (Range) | 70.15<br>(43.0-123.0) | 63.60<br>(47.0-117.0) | 70.30<br>(46.5-122.0) | 69.50<br>(43.0-123.0) |
| BMI | Median (Range) | 24.34<br>(18.03-39.78) | 23.06<br>(19.10-33.50) | 25.34<br>(18.17-43.23) | 24.59<br>(18.03-43.23) |

BMI: body mass index; n : number of patients; SD: standard deviation.

**TS2.** Overall GMT and GMFR for all analysed subvariants.

**TS2. Overall GMT and GMFR for all analysed subvariants.**
|  | Baseline |  | Day 14 |  |  |  |  | 3 months |  |  |  |  | 6 months |  |  |  |  |
| --- | --- | --- | --- | --- | --- | --- | --- | --- | --- | --- | --- | --- | --- | --- | --- | --- | --- |
|  | GMT |  | GMT |  | GMFR |  |  | GMT |  | GMFR |  |  | GMT |  | GMFR |  |  |
|  | Mean | 95% CI | Mean | 95% CI | Mean | 95% CI | p-value | Mean | 95% CI | Mean | 95% CI | p-value | Mean | 95% CI | Mean | 95% CI | p-value |
| **BETA** | 874,17 | 705.201, 1083.626 | 6139,09 | 4951.135, 7612.072 | 6,96 | 5.233, 9.252 | <0.0001 | 4097,39 | 3301.254, 5085.518 | 4,62 | 3.476, 6.149 | <0.0001 | 2236,62 | 1800.007, 2779.146 | 2,54 | 1.906, 3.376 | <0.0001 |
| **DELTA** | 704,53 | 543.274, 913.66 | 4484,67 | 3457.238, 5817.439 | 6,27 | 4.787, 8.215 | <0.0001 | 1771,76 | 1364.578, 2300.433 | 2,49 | 1.900, 3.260 | <0.0001 | 1320,06 | 1015.934, 1715.226 | 1,86 | 1.421, 2.441 | 0,0004 |
| **OMICRON BA.1** | 369,98 | 268.318, 510.164 | 3765,56 | 2730.269, 5193.426 | 9,21 | 5.574, 15.212 | <0.0001 | 1989,17 | 1440.748, 2746.339 | 4,84 | 2.931, 8.005 | <0.0001 | 1814,32 | 1313.071, 2506.919 | 4,42 | 2.676, 7.315 | 0,0001 |
| **OMICRON BA.4/5** | 219,41 | 168.798, 285.186 | 2674,21 | 2055.512, 3479.131 | 11,80 | 8.288, 16.791 | <0.0001 | 1685,38 | 1296.179, 2191.45 | 7,37 | 5.179, 10.495 | <0.0001 | 1687,53 | 1296.932, 2195.765 | 7,40 | 5.193, 10.536 | <0.0001 |
| **OMICRON XBB.1.5** | 51,49 | 40.798, 64.984 | 276,66 | 219.162, 349.238 | 5,22 | 3.966, 6.867 | <0.0001 | 187,20 | 148.209, 236.453 | 3,53 | 2.679, 4.643 | <0.0001 | 185,32 | 146.725, 234.067 | 3,49 | 2.652, 4.596 | <0.0001 |

CI: confidence interval; GMFR: Geometric mean fold rise; GMT: Geometric mean titre.

**TS3.**
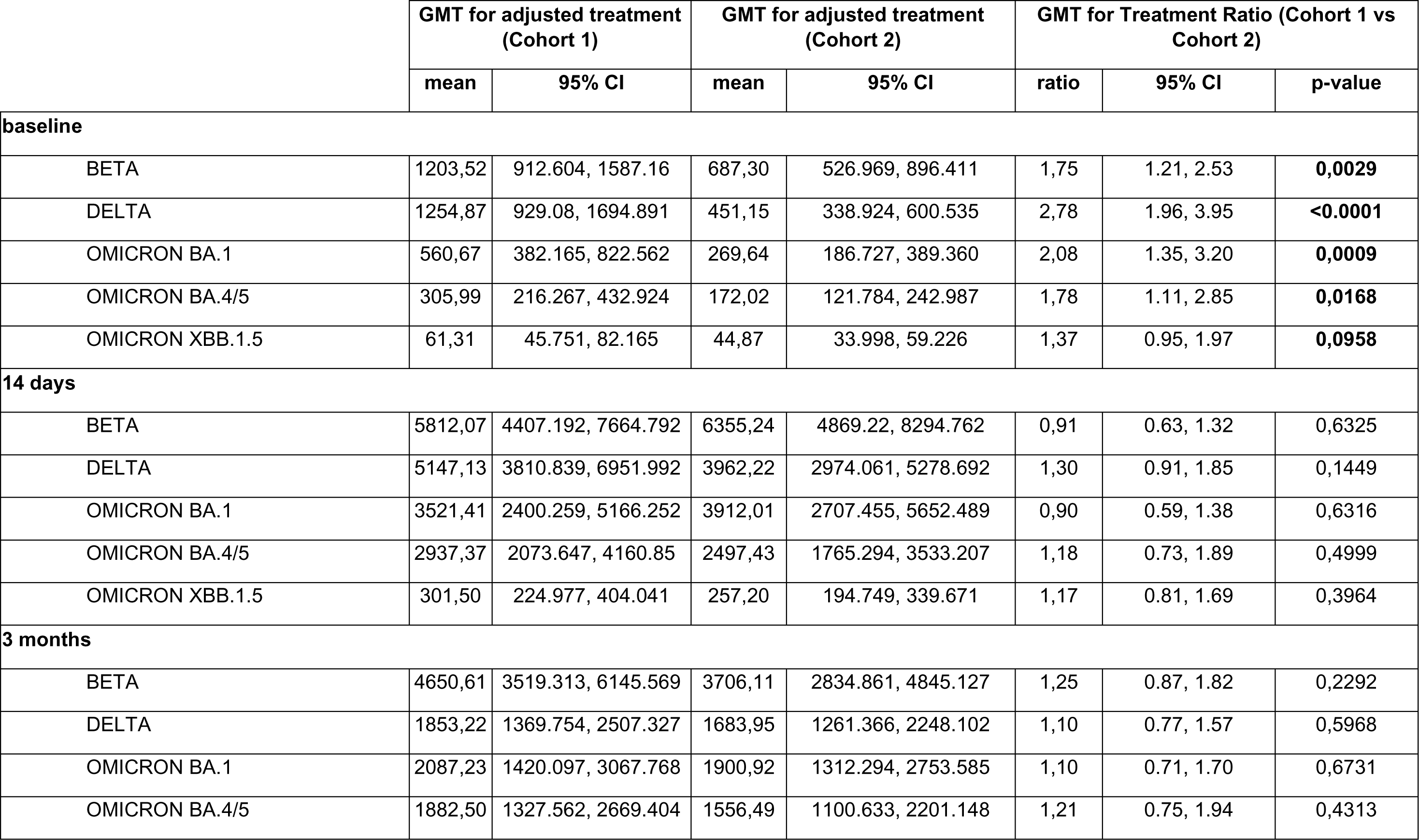
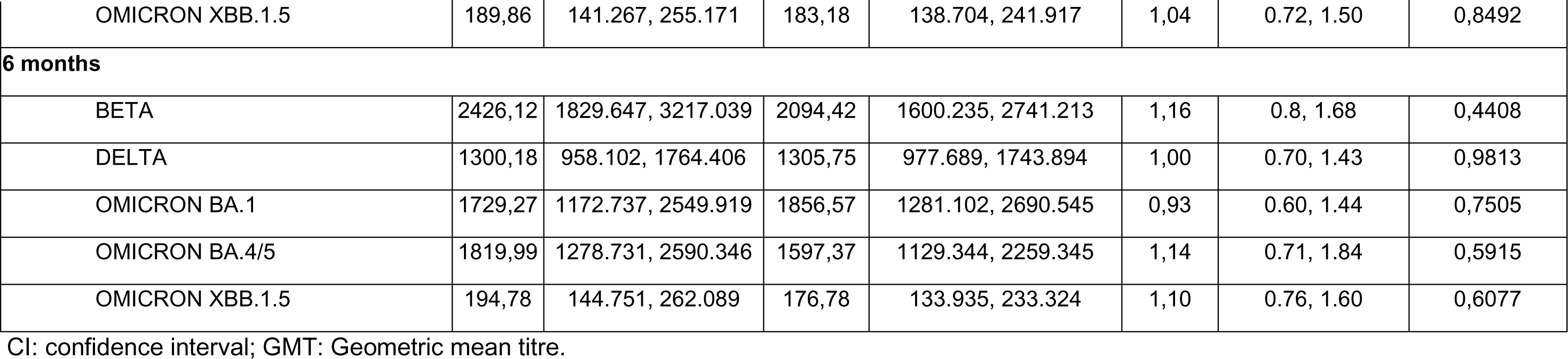
GMT per cohorts for all analysed subvariants.

**Table S4.** Summary of COVID-19 Infections (Safety Population) – Part B.

|  | **Cohort 1:**<br>**PHH-1V + PHH-1V**<br>**(N=106)**<br>**n (%)** | **Cohort 2:**<br>**Comirnaty + PHH-**<br>**1V**<br>**(N=182)**<br>**n (%)** | **Overall**<br>**(N=288)**<br>**n (%)** |
| --- | --- | --- | --- |
| Number of subjects with SARS-CoV-2 infections according to COVID-19 infection criteria throughout the study duration. | 15 (14.2) | 21 (11.5) | 36 (12.5) |
| Responders 95% CI<sup>1</sup> | 8.14, 22.26 | 7.29, 17.10 | 8.91, 16.88 |
| Number of subjects with COVID-19 severe infections through Day 182. | 0 | 0 | 0 |
| Responders 95% CI<sup>1</sup> | NC | NC | NC |
| Number of subjects with hospital admissions associated with COVID-19 through Day 182. | 0 | 0 | 0 |
| Responders 95% CI<sup>1</sup> | NC | NC | NC |
| Number of subjects with ICU admissions associated with COVID-19 through Day 182. | 0 | 0 | 0 |
| Responders 95% CI<sup>1</sup> | NC | NC | NC |
| Number of subjects with deaths associated with COVID-19 through Day 182. | 0 | 0 | 0 |
| Responders 95% CI<sup>1</sup> | NC | NC | NC |

CI: confidence interval; COVID-19: Coronavirus Disease 2019; N: the number of subjects in the population; NC: Not calculable.
<sup>1</sup>A responder is defined as those subjects who meet the criteria. Exact CI for the proportion of responders has been calculated using the Clopper-Pearson method.

